# Revisiting the First Night Effect of Polysomnography Using Wearable EEG Headbands in Older Adults with Sleep Disturbances

**DOI:** 10.64898/2026.11.23.26368802

**Authors:** Anne Chen, Gawon Cho, Ian Murphy, Maamie Asamoah-Mensah, Jarett Talarczyk, Brienne Miner

## Abstract

**Study Objectives:** The first night effect (FNE), where sleep is altered during the first night of polysomnography (PSG), may affect the validity of single-night PSG assessments, yet its magnitude relative to habitual sleep is difficult to characterize using repeated PSG alone. We examined FNE during home-based PSG (HPSG) in older adults with sleep disturbances using multi-night wearable EEG headband (HB) recordings.

**Methods:** Fifty-eight community-dwelling adults aged 55-85 years with sleep disturbances completed up to seven nights of HB monitoring, including one night of simultaneous HPSG. Using HB-derived measures, we compared total sleep time (TST), wake after sleep onset (WASO), sleep efficiency (SE), minutes in N3 sleep, and minutes in REM sleep between HPSG and non-HPSG nights via pairwise comparisons and linear mixed-effects models adjusting for age, sex, and race/ethnicity.

**Results:** Sleep differed significantly between HPSG and non-HPSG nights, with roughly 35 minutes longer WASO, 21 minutes less N3 sleep, and 15% lower SE (*p*’s < .001) during HPSG nights. TST and REM sleep did not differ between HPSG and non-HPSG nights. Findings were consistent when separately comparing nights preceding and following HPSG.

**Conclusions:** In older adults with sleep disturbances, a single night of HPSG was associated with greater wake after sleep onset, less N3 sleep, and lower sleep efficiency than other nights measured with wearable EEG, suggesting that one-night PSG may overstate sleep disruption and understate deep sleep. Multi-night wearable EEG monitoring may help assess the representativeness of single-night PSG measurements, thereby improving the accuracy of sleep measurement interpretation.

**Statement of Significance:** Polysomnography (PSG) is the reference standard for evaluating sleep, yet a single recording night may not reflect how a person usually sleeps. Using a wearable EEG headband worn across multiple nights, we found that older adults with sleep disturbances had more nighttime wakefulness, less deep sleep, and lower sleep efficiency during home PSG than on other nights. These findings suggest that single-night PSG can overstate sleep disruption in this population. Pairing PSG with multi-night wearable monitoring may help clinicians and researchers interpret PSG results more accurately and better judge treatment response.

## INTRODUCTION

Sleep disturbances are highly prevalent among older adults and are associated with impaired daytime functioning, cardiovascular disease, cognitive decline, and reduced quality of life.^1–4^ Because self-reported sleep often disagrees with objectively measured sleep in this population,^5–7^ clinical and research assessment frequently relies on polysomnography (PSG), the reference standard for objective sleep assessment.^8^ However, PSG is typically limited to a single night, and the recording environment, instrumentation, and monitoring procedures may alter sleep, a phenomenon known as the first-night effect (FNE).^9^ Substantial FNE during a single PSG night may produce sleep measurements that are not representative of habitual sleep, potentially reducing their accuracy in informing diagnostic assessments, guiding treatment decisions, and evaluating treatment response in clinical trials.^10^ However, the extent to which FNE affects older adults with sleep disturbances remains insufficiently characterized.^11,12^ To our knowledge, no study has quantified these differences at the EEG level by comparing sleep during a PSG night with sleep on other nights using a less intrusive device in the same individuals. By characterizing differences in sleep between a single PSG night and surrounding nights, this study aims to inform how recording-related changes may influence the interpretation of PSG-derived sleep measures in older adults with sleep disturbances.

Traditionally, FNE has been evaluated by comparing consecutive nights of PSG, which characterizes changes in sleep across repeated recordings but does not establish how sleep during a PSG night differs from sleep measured without PSG instrumentation. Home-based PSG (HPSG) eliminates the laboratory environment^13^ and has been widely validated against conventional laboratory PSG,^14,15^ but retains the instrumentation and monitoring procedures associated with PSG. Less intrusive wearable devices, such as actigraphy, have shown minimal or no FNE^16,17^ but cannot characterize sleep architecture and may be inaccurate when sleep is more fragmented,^18^ as is common in older adults with sleep disturbances. Wearable electroencephalography (EEG) headbands (HBs) offer an opportunity to overcome this limitation by providing EEG-based sleep measurements across multiple nights. Self-applied HBs are less burdensome than PSG systems and have been validated against PSG.^18,19^ Importantly, HBs can be worn simultaneously with HPSG, allowing sleep measurements obtained during a PSG night to be compared with those obtained on other nights without PSG instrumentation. This approach may help characterize HPSG-associated differences in sleep while minimizing the influence of the laboratory environment and repeated PSG instrumentation.

FNE has been well documented in PSG studies and has been associated with longer sleep onset latency (SOL), increased wake after sleep onset (WASO), disrupted REM cycles,^9,20^ reduced total sleep time (TST) and N3 sleep, and lower sleep efficiency (SE).^11,21,22^ However, the magnitude and pattern of these changes appear to vary across populations and recording settings. Studies comparing older adults with insomnia and good sleepers have reported first-night changes in both groups, with findings differing between laboratory and HPSG.^23,24^ One study even reported a reverse FNE, in which participants with insomnia experienced better sleep quality on the first night of laboratory PSG.^25^ Although some evidence suggests that FNE may be more pronounced among older adults, the available literature is limited by small older-adult samples and inconsistent assessment of sleep outcomes.^11^ Whether, and in which direction, sleep during a single PSG night differs from sleep on other nights in this population therefore remains unknown.

To address these gaps, the present study examined PSG-associated differences in sleep among older adults with sleep disturbances using a multi-night, self-applied wearable EEG HB alongside a single night of HPSG. The primary objective was to examine differences in HB-derived sleep outcomes between HPSG night and non-HPSG nights. We hypothesized that relative to non-HPSG nights, the PSG night would be characterized by reduced TST, prolonged WASO, less N3 and REM sleep, and lower SE. By quantifying differences in sleep between a single HPSG night and surrounding HB-only nights, this study aims to inform the interpretation of single-night PSG-derived sleep measures in older adults with sleep disturbances.

## METHODS

### Study Population and Eligibility

A total of 63 community-dwelling older adults aged ≥ 55 years were recruited from the greater New Haven area of Connecticut through flyers distributed at primary care clinics and local senior centers. Participants were eligible if they reported sleep disturbances at least once per week, including symptoms of insomnia (i.e., difficulty initiating or maintaining sleep or early morning awakenings) and/or daytime sleepiness (i.e., frequent dozing or napping). Exclusion criteria included non-English speakers; inability to provide informed consent; residence in an assisted living or extended care facility; or an unplanned hospitalization within the preceding month. Individuals who were otherwise eligible were not excluded based on use of hypnotic medications or a known diagnosis of sleep apnea, insomnia, or restless legs syndrome. Recruitment and data collection took place between January 2022 and December 2023. Details of the study design and data collection protocols have been described previously.^18^

Of the 63 individuals who completed the study, 5 participants were excluded from data analysis (2 had unanalyzable HB data on HPSG night; 2 had fewer than 4 nights of usable HB data from non-HPSG nights; 1 underwent HPSG on the first night of HB recording). Thus, the final analytical sample comprised 58 participants.

### Study Protocol

The study protocol consisted of 7 days of home-based sleep monitoring. Participants wore the HB at home for up to seven nights and completed a single night of simultaneous HPSG mid-week. At baseline (Visit 1), participants provided demographic and clinical information and completed subjective sleep questionnaires. Study staff then introduced participants to the sleep monitoring devices and provided instructions for their use throughout the monitoring period. After two to three nights of acclimation to the HB, participants selected a night for overnight HPSG, which typically fell on the 4^th^ and 5^th^ night. On the selected night, study staff visited participants’ homes to set up the HPSG equipment and initiated the HB recording before leaving. HPSG devices had an automatic timed start selected to occur roughly one hour before the participant’s usual sleep onset. Participants wore the HB and HPSG equipment simultaneously overnight. The following morning, participants removed the HPSG equipment and continued wearing the HB for the remainder of the monitoring period.

### Demographic and Clinical Characteristics

Demographic characteristics included age, sex, race/ethnicity (non-Hispanic white vs. other), education (college graduate vs. other), and occupational status (retired vs. other). Clinical characteristics included body mass index, cognitive impairment (Montreal Cognitive Assessment [MoCA] short form score < 12),^26^ self-reported physician diagnoses of diabetes, coronary artery disease, heart failure, chronic obstructive pulmonary disease, a psychiatric condition (including depression, anxiety, bipolar disorder, post-traumatic stress disorder, or substance use disorder), or a sleep disorder (including sleep apnea, insomnia, or restless legs syndrome). Participants reported prescription and over-the-counter medication use during the preceding 2 weeks. For the total number of medications used, we included prescription medications and over- the-counter use of aspirin, pain medication (e.g., acetaminophen, ibuprofen, naproxen), allergy/cold medicines (e.g., diphenhydramine, cetirizine), or antacids (e.g., tums, famotidine, proton pump inhibitors). Sleep medication use was assessed separately by asking whether participants had taken a prescription or over-the-counter medication or herbal or nutritional supplement as a sleep aid during the preceding 2 weeks.

### Sleep Measures

Self-reported sleep measures included the Pittsburgh Sleep Quality Index (PSQI),^27^ which assesses subjective sleep quality and sleep disturbance over the preceding month; the Epworth Sleepiness Scale (ESS),^28^ which assesses excessive daytime sleepiness; and the Insomnia Severity Index (ISI), which assesses the severity of insomnia symptoms.^29^

Objective sleep measures were obtained using the HB and PSG. The Dreem 2 HB (formerly Dreem, Paris, France; now Beacon Biosignals, Boston, MA, USA) is a wireless, self-administered, noninvasive, FDA Class II medical device. The HB has five dry EEG electrodes, a three-dimensional accelerometer for assessing movement, body position, and respiration, and an infrared pulse oximeter. The EEG electrodes are positioned across the frontal forehead (F7, F8, and FpZ) and posterior occipital region (O1 and O2) of the scalp, generating seven EEG derivations: FpZ–O1, FpZ–O2, FpZ–F7, F8–F7, F7–O1, F8–O2, and FpZ–F8. EEG signals are sampled at 250 Hz and bandpass filtered from 0.4–35 Hz. The device communicates via Bluetooth with a mobile application on a smartphone or tablet, after which data are transferred via Wi-Fi to the study sponsor’s servers. Recordings from each study night were stored on the HB and retrieved at the end of the week-long protocol. Sleep measures were generated using the HB’s proprietary automated sleep-scoring algorithm.

HPSG was conducted in participants’ homes using the NOX-Self Applied system (Nox Medical, Reykjavík, Iceland). The system records standard PSG signals, including electroencephalography (EEG), electrooculography (EOG), electromyography (EMG), airflow, thoracic and abdominal respiratory effort, electrocardiography (ECG), and oxygen saturation. A trained sleep technologist at Sleep Strategies, Inc. manually scored PSG recordings.

The HB and HPSG provided estimates of TST (total time asleep between sleep onset and final awakening), WASO (total time awake between sleep onset and final awakening), SOL (time from attempted sleep onset to sleep onset), SE (ratio of TST to time in bed), and time spent in light sleep (N1 and N2), deep sleep (N3), and rapid eye movement (REM) sleep.

### Statistical Analysis

To evaluate HPSG-associated FNE, we assessed differences between sleep on HPSG versus non-HPSG nights. HB-derived sleep measures from the HPSG night were compared with the average of sleep measures derived from all non-HPSG nights. Because HB recordings on HPSG nights were initiated at a different time than participants’ usual bedtime, manually scored lights-off and light-on times from polysomnography were used to adjust the HB analysis period. Paired-samples t-tests were used to compare sleep outcomes, with Wilcoxon signed-rank tests used for outcomes that did not meet the normality assumption. Effect sizes were calculated to characterize the magnitude of observed differences. Cohen’s *d* was reported for paired-samples t-tests, and rank-biserial correlation was reported for the Wilcoxon tests. To account for repeated sleep measurements within participants and differences in the number of available recordings across participants, linear mixed-effects models with participant-specific random intercepts were fitted to examine differences in sleep outcomes between HPSG and non-HPSG nights. Models were additionally adjusted for age, sex, and race/ethnicity.

In secondary analyses, we examined sleep outcomes on the nights immediately preceding and following HPSG separately to assess whether the observed differences between HPSG and non-HPSG nights were influenced by potential post-HPSG recovery effects. Linear mixed-effects models were fitted with night category (pre-HPSG, HPSG, and post-HPSG) as a three-level fixed effect. Overall differences across night categories were assessed using Type III F-tests with Satterthwaite’s approximation for denominator degrees of freedom, followed by pairwise comparisons of pre-HPSG and post-HPSG nights with the HPSG night. The same sleep outcomes and covariate adjustments used in the primary analysis were applied.

Although the HB is a minimally burdensome wearable device, initial adjustment to wearing the device could influence sleep during the first few nights of monitoring. Therefore, we examined whether sleep outcomes measured by the HB changed over successive nights of use, independent of HPSG exposure, to explore potential FNE or acclimation to the HB device. Analyses were restricted to non-HPSG nights because HB data on the night of PSG could be abnormal due to FNE. Linear mixed-effects models were fitted separately for each sleep outcome, with night of HB use modeled as a categorical fixed effect and adjusted for covariates. Overall differences across nights were assessed using Type III F-tests with Satterthwaite’s approximation for degrees of freedom. Pairwise comparisons of estimated marginal means were performed with Holm adjustment for multiple comparisons.

## RESULTS

The final analytical sample included 58 participants with a mean age of 76.6 ± 6.6 years. Thirty-six (63.2%) were women, forty-four (78.6%) were non-Hispanic White, fifty (87.7%) were college graduates, and twelve (21.1%) had cognitive impairment. Participants averaged 4.22 ± 1.4 nights of HB use before undergoing HPSG and 1.36 ± 1.2 nights after HPSG. Self-reported sleep disturbances included sleep apnea (24.1%), insomnia (12.1%), and restless leg syndrome (3.4%). Additional demographic and clinical characteristics of the study population are presented in Table 1.

**Table 1.** Demographic and clinical characteristics of the sample.

| **Characteristics** | **N=58**<br>Mean (SD) or n (%) |
|---|---|
| Age, *years* | 76.6 (6.6) |
| Female | 37 (63.8%) |
| Non-Hispanic White | 45 (78.9%) |
| College graduate | 51 (87.9%) |
| Retired | 39 (67.2 %) |
| Body mass index | 28.3 (7.2) |
| Cognitive impairment<sup>a</sup> | 12 (21.1%) |
| History of diabetes<sup>b</sup> | 9 (15.5%) |
| History of coronary artery disease<sup>b</sup> | 5 (8.6%) |
| History of heart failure<sup>b</sup> | 4 (6.9%) |
| History of chronic obstructive pulmonary disease<sup>b</sup> | 8 (13.8%) |
| History of a psychiatric disorder<sup>c</sup> | 23 (39.7%) |
| History of sleep disorder<sup>b</sup> | 20 (34.5%) |
| Sleep Apnea<sup>d</sup> | 14 (24.1%) |
| Insomnia | 7 (12.1%) |
| Restless legs syndrome | 2 (3.4%) |
| Medication count<sup>f</sup> | 7.2 (4.7) |
| Reported use of a sleep aid in the past two weeks<sup>g</sup> | 18 (31.6%) |
| Pittsburgh Sleep Quality Index score | 9.4 (3.9) |
| Epworth Sleepiness Scale score | 8.0 (5.1) |
| Insomnia Severity Index score | 13.0 (4.6) |
| Total sleep time, *minutes* <sup>h</sup> | 370.5 (92.5) |
| Wake after sleep onset, *minutes*<sup>h</sup> | 84.5 (62.0) |
| Apnea hypopnea index, *events/hour*<sup>h</sup> | 19.3 (15.3) |
| Periodic limb movement index, *events/hour* <sup>h</sup> | 7.7 (14.7) |

<sup>a</sup> Score <12 on the Montreal Cognitive Assessment short form.
<sup>b</sup> Self-reported physician diagnosis
<sup>c</sup> Self-reported physician diagnosis of at least one of the following: depression (13), anxiety (17), bipolar disorder (1), post-traumatic stress disorder (3), substance use disorders (1).
<sup>d</sup> 2 participants used continuous positive airway pressure therapy during the study.
<sup>f</sup> Total number of medications taken in the past two weeks, including prescription medications and over-the-counter use of aspirin, pain medication (e.g., acetaminophen, ibuprofen, naproxen), allergy/cold medicines, or antacids (e.g., tums, famotidine, proton pump inhibitor); does not include use of bowel regimens, eyedrops, topical lotions, nasal sprays, multivitamins or probiotics.
<sup>g</sup> Inquired by asking, “In the past two weeks, have you taken a sleep aide to help you sleep during the night?” and includes prescription or over-the-counter medications or supplements.
<sup>h</sup> From polysomnography

HB-derived sleep outcomes were compared between HPSG and non-HPSG nights. As shown in Table 2, HPSG nights had significantly longer TST, greater WASO, shorter N3 sleep duration, and lower SE, whereas REM duration did not differ significantly. The largest standardized difference was observed for SE (Cohen’s *d* = −1.38), followed by N3 sleep (rank-biserial correlation = −0.65) and WASO (*d* = 0.53), while the difference in TST was smaller (*d* = 0.27). Results from adjusted mixed-effects models were consistent with the paired comparisons. Relative to non-PSG nights, PSG nights were associated with 19 minutes longer TST (*p* = 0.04), 35 minutes longer WASO (*p* < .001), 21 minutes less N3 sleep (*p* < .001), and 15 percentage points lower SE (*p* < .001). REM duration remained similar between HPSG and non-HPSG nights.

**Table 2.** Sleep outcomes on PSG vs. Non-PSG Nights.

| **Outcome** | **PSG (SD)** | **Non-PSG (SD)** | **Paired effect size,<br>*p*** | **Unadjusted β<br>(95% CI)** | **Adjusted β<br>(95% CI)** |
|---|---|---|---|---|---|
| TST (hr) | 6.45 (1.6) | 6.12 (1.0) | *d* = 0.27, .042\* | 0.33 (0.02, 0.64) | 0.32 (0.01, 0.63) |
| WASO (min) | 106.11 (74.8) | 71.45 (38.8) | *d* = 0.53, <.001\* | 34.60 (21.18, 48.01) | 35.06 (21.48, 48.64) |
| N3 (min) | 27.70 (29.8) | 48.89 (27.0) | *r* = −0.65, <.001\* | −21.18 (−27.92, −14.43) | −21.24 (−28.05, −14.43) |
| REM (min) | 87.59 (47.7) | 91.35 (39.0) | *d* = −0.09, .502 | −3.75 (−13.98, 6.47) | −3.45 (−13.59, 6.69) |
| SE (%) | 59.93 (12.8) | 74.92 (9.4) | *d* = −1.38, <.001\* | −15.03 (−17.59, −12.48) | −14.99 (−17.57, −12.42) |

**Notes:** Paired effect sizes are Cohen’s *d* for TST, WASO, REM, and SE and rank-biserial correlation (*r*) for N3. Mixed-effects model estimates (β) represent the difference between PSG and non-PSG nights (PSG − non-PSG). Unadjusted models included PSG status and a participant-level random intercept. Adjusted models additionally included age, sex, and race/ethnicity. Confidence intervals are 95%.
\* Denotes statistical significance at the level of *p*<.0.05.

When comparing pre-HPSG, HPSG, and post-HPSG nights, overall differences across the three periods were observed for WASO (*F*(2, 331.7) = 13.19, *p* < .001), N3 sleep (*F*(2, 326.2) = 19.30, *p* < .001), and SE (*F*(2, 327.3) = 66.0, *p* < .001), but not for TST and REM duration (Table 3). Pairwise comparisons showed that both pre-HPSG nights and post-HPSG nights had shorter WASO, longer N3 sleep, and higher SE relative to HPSG nights. For all sleep outcomes, there was no significant difference between Pre-PSG and Post-PSG nights.

**Table 3.** Sleep outcomes on Pre-PSG, PSG, and Post-PSG nights.

| **Sleep outcome** | **Pre-PSG, EMM (95% CI)** | **PSG, EMM (95% CI)** | **Post-PSG, EMM (95% CI)** | **Omnibus *p*** | **PSG vs. Pre-PSG, Δ (adjusted *p*)** | **PSG vs. Post-PSG, Δ (adjusted *p*)** |
|---|---|---|---|---|---|---|
| TST (hr) | 6.13 (5.84–6.43) | 6.45 (6.07–6.84) | 6.11 (5.74–6.48) | .110 | 0.32 (.136) | 0.35 (.157) |
| WASO (min) | 72.8 (61.7–83.9) | 106.1 (90.8–121.5) | 67.6 (53.1–82.2) | <.001* | 33.3 (<.001)* | 38.5 (<.001)* |
| N3 (min) | 49.6 (42.6–56.7) | 27.7 (18.9–36.5) | 46.5 (38.1–55.0) | <.001* | −21.9 (<.001)* | −18.8 (<.001)* |
| REM (min) | 91.1 (80.8–101) | 87.6 (74.5–101) | 92.1 (79.5–101) | .757 | −3.5 (1.00) | −4.5 (1.00) |
| SE (%) | 74.7 (72.2–77.3) | 59.9 (56.7–63.2) | 75.7 (72.6–78.7) | <.001* | −14.8 (<.001)* | −15.7 (<.001)* |

**Note:** Values are estimated marginal means (EMMs) with 95% confidence intervals from linear mixed-effects models with a participant-level random intercept. Pairwise differences represent PSG minus Pre-PSG and PSG minus Post-PSG, respectively. *P*-values for pairwise comparisons were adjusted using the Holm method. Pre-PSG versus Post-PSG comparisons were not statistically significant for any outcome (all adjusted *p* > .36), indicating no evidence of a post-PSG recovery effect.
\* Denotes statistical significance at the level of *p*<.0.05.

When comparing successive non-HPSG nights, no significant differences in sleep outcomes were observed across nights of HB use (Table 4).

**Table 4.**
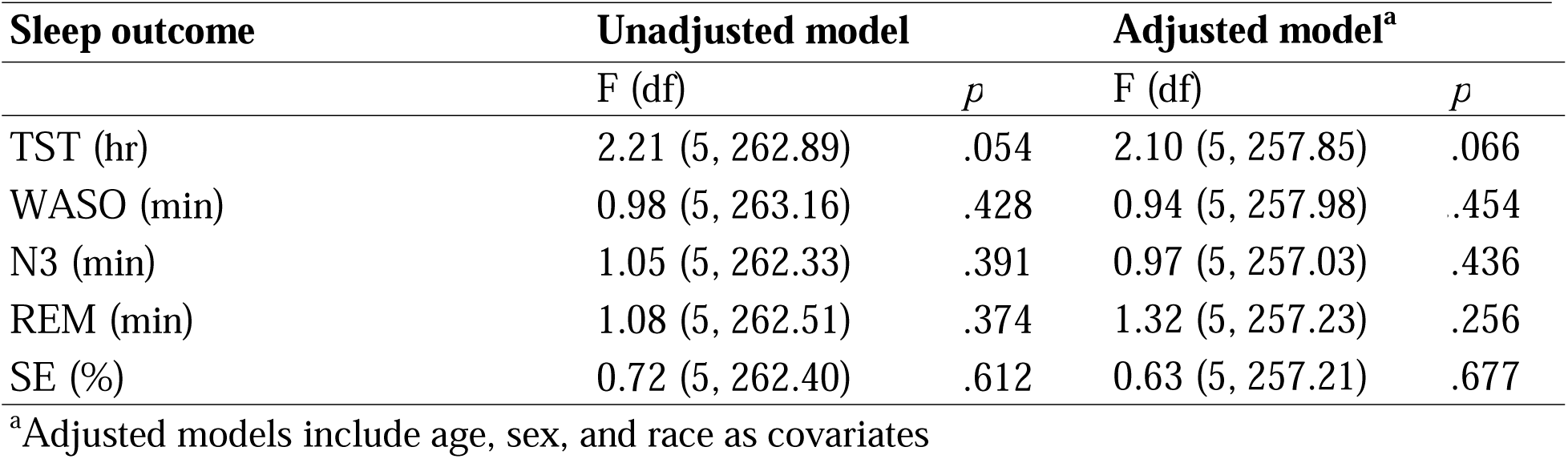
Differences in sleep outcomes across non-PSG nights.

## DISCUSSION

In this study of 58 older adults with self-reported sleep disturbances, we identified FNE associated with a single HPSG night within a multi-night wearable EEG HB monitoring period. By leveraging repeated sleep measurements from the same individuals, we made within-person comparisons of sleep between HPSG nights and non-HPSG nights. Compared with non-HPSG nights, the HPSG night had, on average, 35 more minutes of WASO, 21 fewer minutes of N3 sleep, and 15% lower SE. These differences were still observed when the HPSG night was examined in relation to the immediately preceding and following nights. More importantly, we found no significant overall changes in sleep outcomes across successive non-HPSG nights. To our knowledge, this is the first study to evaluate HPSG-associated FNE by applying a multi-night, minimally invasive wearable EEG monitoring HB. Unlike traditional FNE designs that compare the first PSG night with one or more subsequent PSG nights, our design allowed us to characterize sleep on the HPSG night relative to less instrumented nights in the home environment. The results quantify FNE in a home-based environment and highlight the potential value of wearable EEG devices for obtaining repeated measures of sleep at home while accounting for differences associated with polysomnography.

Our findings extend the FNE literature to older adults with sleep disturbances, a population that has received limited attention. Classically, FNE has been characterized by longer SOL, more WASO, disrupted REM sleep, shorter TST and N3 sleep, and lower SE on the first night of polysomnography, mostly in healthy middle-aged sleepers or people with insomnia.^11,22,30^ Studies in older adults have involved smaller samples, younger participants, or both: Coates et al.^23^ compared 12 older adults with insomnia with 12 good sleepers (maximum age 60), and Edinger et al.^24^ and Riedel et al.^31^ studied larger samples of adults with insomnia (mean age 68). Our participants were older (mean age 76.6 years) and had a broader range of sleep complaints, including sleep apnea, insomnia, and restless legs syndrome.

Consistent with previous studies, we observed longer WASO, lower SE, and shorter N3 sleep on the HPSG night, three commonly reported manifestations of FNE in laboratory-based PSG studies. Despite limited evidence regarding HPSG-associated changes in WASO among older adults with sleep disturbances, our observed 35-minute increase was comparable to the 52- minute difference reported between the first and second nights of HPSG in a small sample of adults over 80 years old.^32^ In contrast, a study of adults with insomnia with a maximum age of 60 years reported a smaller, nonsignificant difference of approximately 16 minutes between the first and subsequent HPSG nights.^23^ Differences in participant age may partly explain the variation in observed WASO differences across studies. SE was approximately 15 percentage points lower on the PSG night than on non-PSG nights, exceeding the 5 percentage point reduction reported for the first night in a meta-analysis of healthy older adults with no sleep disturbances undergoing two consecutive nights of PSG.^11^ Similarly, Riedel et al. reported an 8 percentage point reduction in SE and a 3 percentage point reduction in N3 sleep on the first night among older adults with insomnia undergoing laboratory PSG.^15^ In parallel with our study, Edinger et al. also observed roughly 20 minutes less N3 sleep on the first night of both laboratory and HPSG in insomniacs, but a significant reduction in SE was evident only during laboratory monitoring.^27^ The larger differences in sleep outcomes observed in our study may partly reflect our use of less intrusive, multi-night wearable EEG recordings to establish a reference closer to habitual sleep, rather than comparisons between consecutive PSG nights. This approach provides a complementary perspective on instrumentation-associated FNE during HPSG.

In contrast to several classic manifestations of FNE, we did not find reduced TST on the HPSG night. In our primary analysis, TST was approximately 19 minutes longer on the PSG night than on non-HPSG nights. However, comparisons across pre-HPSG, HPSG, and post-HPSG periods did not identify overall significant differences. This finding differs from studies reporting shorter TST during initial PSG recordings, particularly in laboratory settings.^30,32–34^ One possible explanation is our home-based protocol, in which HPSG equipment was applied several hours before bedtime, potentially extending time in bed and leading to longer sleep duration. Thus, although sleep continuity and architecture were altered, these changes did not translate into shorter TST in our study.

Similarly, REM duration did not differ significantly between HPSG and non-HPSG nights. However, comparisons with previous studies are complicated by differences in the REM parameters examined. Earlier FNE studies in older adults have more commonly reported changes in REM latency or the distribution of REM sleep rather than total REM duration.^23,32^ Furthermore, the absence of a difference in REM duration should be interpreted cautiously given the potential limitations of the wearable EEG HB in measuring sleep stages, particularly REM sleep. Taken together, these findings suggest that FNE may manifest differently across sleep parameters and that observed changes in TST and REM may depend on both recording protocols and measurement methods.

Importantly, our study demonstrates that FNE can occur during HPSG despite monitoring in a familiar environment. Although conducting PSG at home has been proposed to eliminate or mitigate FNE,^34,35^ previous studies suggest that the home environment may attenuate, but does not necessarily eliminate, these effects. Studies in healthy adults found no significant differences in FNE between laboratory and HPSG studies, suggesting that a familiar environment alone is insufficient to eliminate PSG-associated changes in sleep.^11^ In contrast, another study involving healthy sleepers and individuals with insomnia found more pronounced FNE during laboratory PSG, whereas HPSG-associated changes were largely limited to reduced N3 sleep, suggesting some attenuation of FNE in the home setting.^24^ Our findings indicate that sleep changes exist during HPSG monitoring when compared with HB-measured sleep. A familiar environment may reduce anxiety and disruption associated with laboratory monitoring, but PSG instrumentation and awareness of being monitored may continue to influence sleep. Because these factors cannot be isolated in our study, the observed changes should be interpreted as HPSG-associated rather than attributable to instrumentation alone.

A key finding of our study was the absence of systematic changes in sleep outcomes across non-HPSG nights, supporting the potential of wearable EEG HBs to characterize near- habitual sleep over multiple nights. Unlike conventional FNE studies, which generally compared consecutive PSG nights, our approach allowed sleep on a HPSG night to be compared with multiple nights of objective sleep measurements obtained without HPSG instrumentation. Comparisons of pre- and post-HPSG nights showed no significant differences, providing no evidence of a sustained change in sleep following HPSG. By using the same HB across HPSG and non-HPSG nights, our study provided a consistent method of sleep assessment while only introducing additional HPSG instrumentation on a single night. This approach offers a complementary framework for evaluating HPSG-associated changes against repeated measurements of sleep under less intrusive monitoring conditions. Although the absence of changes in sleep pattern in HB-only nights does not necessarily exclude an initial adaptation effect or other measurement-related influences, multi-night wearable EEG monitoring may help contextualize single-night HPSG measurements relative to an individual’s near-habitual sleep.

This study has several strengths. First, the multi-night study design provided repeated objective measurements of sleep before and after HPSG, allowing us to characterize sleep variability across non-HPSG nights and evaluate HPSG-associated changes relative to multiple nights of sleep rather than relying solely on comparisons between consecutive PSG recordings. Second, the use of a self-applied, minimally intrusive wearable EEG HB allowed us to compare HPSG-night sleep with sleep measured without the additional instrumentation required for PSG. This approach may provide a closer approximation of habitual sleep than protocols involving multiple nights of PSG. While we cannot exclude the possibility that wearing the headband itself influenced sleep, the absence of changes in sleep outcomes across non-HPSG nights suggests that any such effects were minimal.

Several limitations should be considered. First, not all participants completed the 7-night protocol, so nights later in the protocol had fewer observations and less precise estimates. The number of nights before and after HPSG were also asymmetric (4.22 nights before vs. 1.36 nights after). Second, the HB was worn simultaneously with HPSG; therefore, we cannot isolate the effects of HPSG instrumentation from the additional burden of wearing both devices. However, because the HB was also worn on non-HPSG nights and sleep outcomes did not show systematic changes across these nights, the HB alone is unlikely to explain the observed or increased differences. Third, while the HB has good agreement with HPSG for measures of TST, WASO, and SE, agreement is lower for sleep-stage classification.^19,36^ Because our analyses compared HB-derived measurements across all nights rather than directly comparing HB measurements with PSG, differences between the two measurement methods are not likely to explain our findings. In addition, SOL was not available for the PSG night due to protocol- related factors, precluding its evaluation as an FNE outcome. Lastly, the sample was predominantly non-Hispanic White and college-educated, which may limit generalizability to more diverse populations and to older adults without sleep complaints.

In summary, we found that sleep measurements obtained during a single night of HPSG may differ meaningfully from an individual’s sleep across other nights, even when monitoring occurs in a familiar home environment. The magnitude of the changes in sleep pattern was greater than that reported in several previous studies of older adults, potentially due to the use of multi-night, less intrusive EEG monitoring to characterize sleep outside PSG recordings. These findings highlight the importance of considering PSG-associated sleep changes when interpreting single-night sleep measurements in older adults with sleep disturbances. Although wearable EEG is not sufficient to replace PSG for comprehensive sleep assessment, it may complement PSG by providing additional information about an individual’s sleep patterns beyond a single monitored night.

## Data Availability

All data produced in the present study are available upon reasonable request to the authors

## FUNDING

This work was supported by the National Institute on Aging (grant numbers R03AG073991, K76AG074905) and the Claude D. Pepper Older Americans Independence Center at Yale School of Medicine (P30AG021342). This manuscript is the result of funding by the National Institutes of Health (NIH). It is subject to the NIH public access policy. Through acceptance of this federal funding, NIH has been given the right to make this manuscript publicly available in PubMed Central upon the Official Date of Publication, as defined by the NIH.

## DISCLOSURES

Financial disclosures: None. Specifically, no funding was provided by the manufacturer of the devices tested in this study.

Non-financial disclosures: None. Specifically, the authors report no conflicts of interest related to this work.

## Notes

### Competing Interest Statement

The authors have declared no competing interest.

### Author Declarations

The Yale University Human Investigation Committee approved the study protocol, and all participants provided written informed consent.

## REFERENCES

1. Miner B, Kryger MH. Sleep in the Aging Population. Sleep Med Clin. 2017;12(1):31–38. doi:10.1016/j.jsmc.2016.10.008

2. Kavousi P, Mali E, Seifhashemi N, Souri M, Pakravan L, Khalili F. Worldwide Prevalence of Poor Sleep Quality in Older Adults: A Systematic Review and Meta-Analysis. Iran J Psychiatry. 2025;20(2):265–280. doi:10.18502/ijps.v20i2.18207

3. da Silva AA, de Mello RGB, Schaan CW, Fuchs FD, Redline S, Fuchs SC. Sleep duration and mortality in the elderly: a systematic review with meta-analysis. BMJ Open. 2016;6(2):e008119. doi:10.1136/bmjopen-2015-008119

4. Mc Carthy CE. Sleep Disturbance, Sleep Disorders and Co-Morbidities in the Care of the Older Person. Med Sci (Basel). 2021;9(2):31. doi:10.3390/medsci9020031

5. Scarlett S, Nolan HN, Kenny RA, O’Connell MDL. Discrepancies in self-reported and actigraphy-based sleep duration are associated with self-reported insomnia symptoms in community-dwelling older adults. Sleep Health: Journal of the National Sleep Foundation. 2021;7(1):83–92. doi:10.1016/j.sleh.2020.06.003

6. Chen JH, Waite L, Kurina LM, Thisted RA, McClintock M, Lauderdale DS. Insomnia Symptoms and Actigraph-Estimated Sleep Characteristics in a Nationally Representative Sample of Older Adults. J Gerontol A Biol Sci Med Sci. 2015;70(2):185–192. doi:10.1093/gerona/glu144

7. Luyster FS, Choi J, Yeh CH, Imes CC, Johansson AEE, Chasens ER. Screening and Evaluation Tools for Sleep Disorders in Older Adults. Appl Nurs Res. 2015;28(4):334–340. doi:10.1016/j.apnr.2014.12.007

8. Practice Parameters for the Indications for Polysomnography and Related Procedures. Sleep. 1997;20(6):406–422. doi:10.1093/sleep/20.6.406

9. Agnew Jr. HW, Webb WB, Williams RL. The First Night Effect: An Eeg Studyof Sleep. Psychophysiology. 1966;2(3):263–266. doi:10.1111/j.1469-8986.1966.tb02650.x

11. Ding L, Chen B, Dai Y, Li Y. A meta-analysis of the first-night effect in healthy individuals for the full age spectrum. Sleep Medicine. 2022;89:159–165. doi:10.1016/j.sleep.2021.12.007

12. Miner B, Pan Y, Cho G, et al. Insomnia with objective short sleep duration, objective long sleep duration, and mortality in older persons. Sleep Health: Journal of the National Sleep Foundation. 2026;0(0). doi:10.1016/j.sleh.2026.08.013

13. Zhang Y, Chen K, Xu Y, Wu H. Research progress on the first-night effect in polysomnography studies. Sleep Research. 2024;1(1):19–26. doi:10.1002/slp2.12

14. Bruyneel M, Ninane V. Unattended home-based polysomnography for sleep disordered breathing: Current concepts and perspectives. Sleep Medicine Reviews. 2014;18(4):341–347. doi:10.1016/j.smrv.2013.12.002

15. Andrade L, Paiva T. Ambulatory Versus Laboratory Polysomnography in Obstructive Sleep Apnea: Comparative Assessment of Quality, Clinical Efficacy, Treatment Compliance, and Quality of Life. J Clin Sleep Med. 2018;14(8):1323–1331. doi:10.5664/jcsm.7264

16. McCall C, McCall WV. Objective vs. Subjective Measurements of Sleep in Depressed Insomniacs: First Night Effect or Reverse First Night Effect? J Clin Sleep Med. 2012;8(1):59–65. doi:10.5664/jcsm.1664

17. Withrow D, Roth T, Koshorek G, Roehrs T. Relation between ambulatory actigraphy and laboratory polysomnography in insomnia practice and research. J Sleep Res. 2019;28(4):e12854. doi:10.1111/jsr.12854

18. Miner B, Chen A, Pan Y, et al. Performance of an electroencephalography-measuring headband or actigraphy compared with polysomnography in older adults with sleep disturbances. Sleep. 2026;49(5):zsag053. doi:10.1093/sleep/zsag053

19. G Ravindran KK, della Monica C, Atzori G, et al. Evaluation of Dreem headband for sleep staging and EEG spectral analysis in people living with Alzheimer’s disease and older adults. Sleep. 2025;48(8):zsaf122. doi:10.1093/sleep/zsaf122

20. Webb WB, Campbell SS. The first night effect revisited with age as a variable. Waking Sleeping. 1979;3(4):319–324.

21. Hof Zum Berge A, Kellmann M, Jakowski S. The First-Night Effect in Elite Sports: An Initial Glance on Polysomnography in Home-Based Settings. Front Psychol. 2021;12:641451. doi:10.3389/fpsyg.2021.641451

22. Sprajcer M, Gupta C, Roach G, Sargent C. Can we put the first night effect to bed? An analysis based on a large sample of healthy adults. Chronobiol Int. 2022;39(12):1567–1573. doi:10.1080/07420528.2022.2133611

23. Coates TJ, George JM, Killen JD, Marchini E, Hamilton S, Thorensen CE. First night effects in good sleepers and sleep-maintenance insomniacs when recorded at home. Sleep. 1981;4(3):293–298. doi:10.1093/sleep/4.3.293

24. Edinger JD, Fins AI, Sullivan RJ Jr, et al. Sleep in the Laboratory and Sleep at Home: Comparisons of Older Insomniacs and Normal Sleepers. Sleep. 1997;20(12):1119–1126. doi:10.1093/sleep/20.12.1119

25. Byun JH, Kim KT, Moon H jin, Motamedi GK, Cho YW. The first night effect during polysomnography, and patients’ estimates of sleep quality. Psychiatry Research. 2019;274:27–29. doi:10.1016/j.psychres.2019.02.011

26. Horton DK, Hynan LS, Lacritz LH, Rossetti HC, Weiner MF, Cullum CM. An Abbreviated Montreal Cognitive Assessment (MoCA) for Dementia Screening. The Clinical Neuropsychologist. 2015;29(4):413–425. doi:10.1080/13854046.2015.1043349

27. Buysse DJ, Reynolds CF, Monk TH, Berman SR, Kupfer DJ. The Pittsburgh Sleep Quality Index: a new instrument for psychiatric practice and research. Psychiatry Res. 1989;28(2):193–213. doi:10.1016/0165-1781(89)90047-4

28. Johns MW. A New Method for Measuring Daytime Sleepiness: The Epworth Sleepiness Scale. Sleep. 1991;14(6):540–545. doi:10.1093/sleep/14.6.540

29. Bastien CH, Vallières A, Morin CM. Validation of the Insomnia Severity Index as an outcome measure for insomnia research. Sleep Medicine. 2001;2(4):297–307. doi:10.1016/S1389-9457(00)00065-4

30. Newell J, Mairesse O, Verbanck P, Neu D. Is a one-night stay in the lab really enough to conclude? First-night effect and night-to-night variability in polysomnographic recordings among different clinical population samples. Psychiatry Research. 2012;200(2):795–801. doi:10.1016/j.psychres.2012.07.045

31. Riedel BW, Winfield CF, Lichstein KL. First night effect and reverse first night effect in older adults with primary insomnia: does anxiety play a role? Sleep Medicine. 2001;2(2):125–133. doi:10.1016/S1389-9457(00)00054-X

32. Wauquier A, van Sweden B, Kerkhof GA, Kamphuisen HAC. Ambulatory first night sleep effect recording in the elderly. Behavioural Brain Research. 1991;42(1):7–11. doi:10.1016/S0166-4328(05)80034-8

33. Iber C, Redline S, Gilpin AMK, et al. Polysomnography Performed in the Unattended Home Versus the Attended Laboratory Setting—Sleep Heart Health Study Methodology. Sleep. 2004;27(3):536–540. doi:10.1093/sleep/27.3.536

34. Bruyneel M, Sanida C, Art G, et al. Sleep efficiency during sleep studies: results of a prospective study comparing home-based and in-hospital polysomnography. Journal of Sleep Research. 2011;20(1pt2):201–206. doi:10.1111/j.1365-2869.2010.00859.x

35. Mamelak A, Hobson JA. Nightcap: a home-based sleep monitoring system. Sleep. 1989;12(2):157–166. doi:10.1093/sleep/12.2.157

36. Arnal PJ, Thorey V, Debellemaniere E, et al. The Dreem Headband compared to polysomnography for electroencephalographic signal acquisition and sleep staging. Sleep. 2020;43(11):zsaa097. doi:10.1093/sleep/zsaa097

